# Race, Socioeconomic Deprivation, and Hospitalization for COVID-19 in English participants of a National Biobank

**DOI:** 10.1101/2020.04.27.20082107

**Authors:** Aniruddh P. Patel, Manish D. Paranjpe, Nina P. Kathiresan, Manuel A. Rivas, Amit V. Khera

**Affiliations:** Center for Genomic Medicine and Division of Cardiology, Department of Medicine, Massachusetts General Hospital, Boston, MA; Harvard Medical School, Boston, MA; Milton Academy, Milton, MA; Department of Biomedical Data Science, Stanford University, Stanford, CA

**Author notes:** Dr. Patel and Mr. Paranjpe contributed equally. Please address correspondence to: Amit V. Khera, MD, MSc, Massachusetts General Hospital, 185 Cambridge Street | CPZN 6.256, Boston, MA 02114, Twitter: @amitvkhera.

## Abstract

Preliminary reports suggest that the Coronavirus Disease 2019 (COVIDâ^’19) pandemic has led to disproportionate morbidity and mortality among historically disadvantaged populations. The extent to which these disparities are related to socioeconomic versus biologic factors is largely unknown. We investigate the racial and socioeconomic associations of COVIDâ^’19 hospitalization among 418,794 participants of the UK Biobank, of whom 549 (0.13%) had been hospitalized. Both black participants (odds ratio 3.4; 95%CI 2.4â^’4.9) and Asian participants (odds ratio 2.1; 95%CI 1.5â^’3.2) were at substantially increased risk as compared to white participants. We further observed a striking gradient in COVIDâ^’19 hospitalization rates according to the Townsend Deprivation Index â^’ a composite measure of socioeconomic deprivation â^’ and household income. Adjusting for such factors led to only modest attenuation of the increased risk in black participants, adjusted odds ratio 3.1 (95%CI 2.0â^’4.8). These observations confirm and extend earlier preliminary and lay press reports of higher morbidity in non-white individuals in the context of a large population of participants in a national biobank. The extent to which this increased risk relates to variation in pre-existing comorbidities, differences in testing or hospitalization patterns, or additional disparities in social determinants of health warrants further study.

## INTRODUCTION

The Coronavirus Disease 2019 (COVID-19) pandemic has led to substantial morbidity and mortality globally, but initial reports suggest historically disadvantaged populations may be most afflicted. For example, a preliminary report from the United States Centers for Disease Control and Prevention indicated that 33% of hospitalized patients were black as compared to 18% of the U.S. population.^1^ Within the densely populated and largely Hispanic suburb of Chelsea, Massachusetts, serologic tests suggested that up to 32% of individuals had been exposed.^2^

The extent to which these disparities are related to socioeconomic versus biologic factors is largely unknown.

## METHODS

The UK Biobank is a prospective cohort study that recruited over 500,000 middle-aged individuals between the years 2006 and 2010, allowing for linkage of extensive baseline, genetic and clinical data.^3^ Recently, COVID-19 testing results for a subset of participants in England were made available.^4^ Individuals were excluded from this analysis if they had enrolled outside England, died prior to 2019, or had an outpatient test positive for COVID-19, in whom the clinical trajectory was uncertain. Additional details are provided in the Supplementary Methods.

## RESULTS

To quantify the relationship between race and hospitalization for COVID-19, 418,794 UK Biobank participants were studied. Mean age at last date of follow-up was 65.8 years, and 188,914 (45%) were male. 400,438 (95.6%) self-reported as white, 10,642 (2.5%) as Asian, and 7,412 (1.8%) as black. 549 (0.13%) individuals were hospitalized with a positive test for COVID-19, detected at 17 assessment centers across England.

COVID-19 hospitalization was noted in 32 of 7,714 (0.4%) black participants, 28 of 10,614 (0.2%) Asian participants, and 489 of 400,438 (0.1%) white participants, with results largely consistent across English regions (**Figure, Panel A**). In a logistic regression model adjusted for age, sex, and geographic region, both black participants (odds ratio 3.4; 95%CI 2.4–4.9) and Asian participants (odds ratio 2.1; 95%CI 1.5–3.2) were at increased risk as compared to white participants.

**FIGURE:**
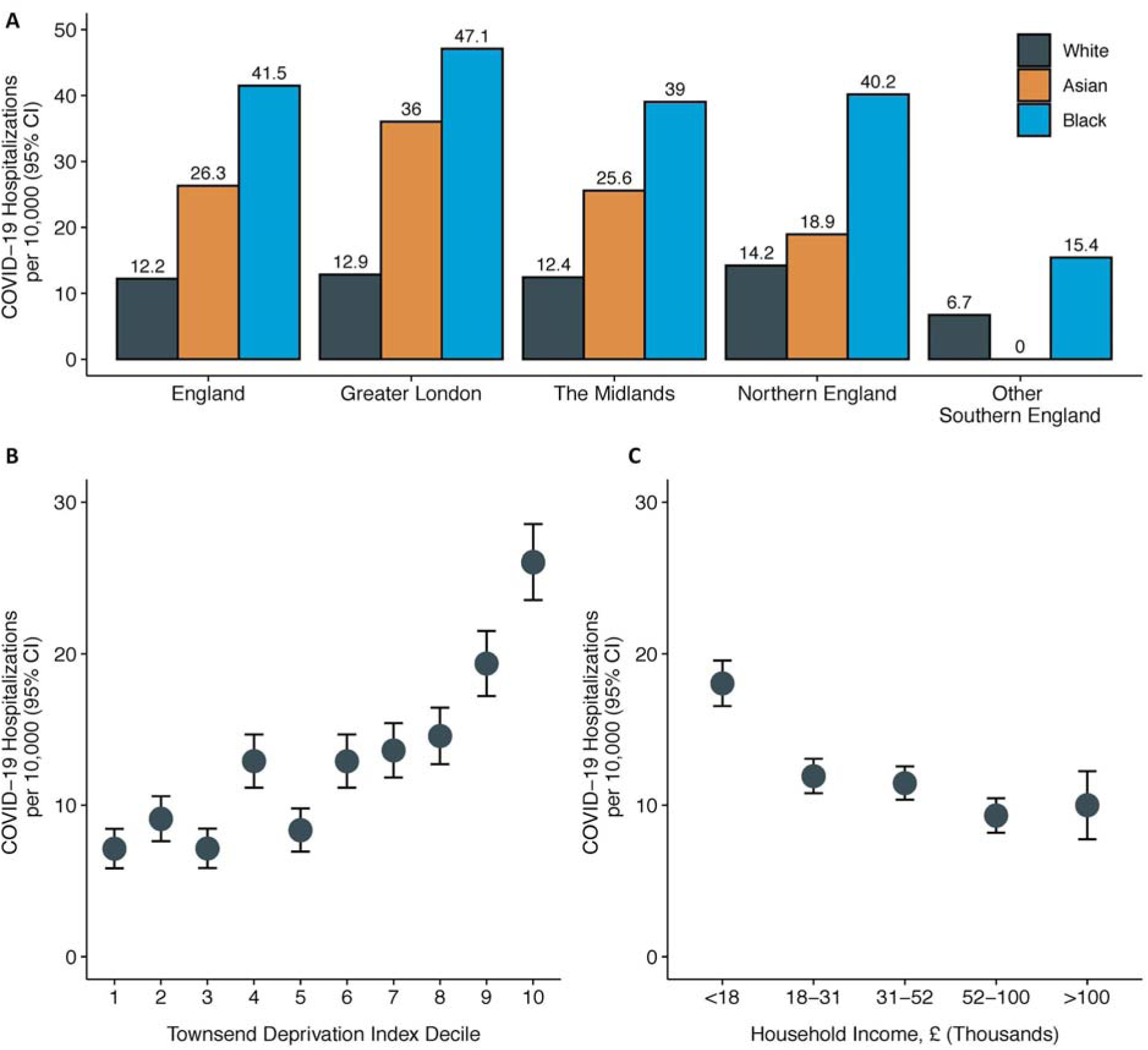
COVID-19 hospitalization rates by race and geographic region, Townsend Deprivation Index, and household income **A:** COVID-19 hospitalizations per 10,000 individuals stratified by self-reported race and geographic region grouped based on socio-demographic similarities. In all regions, significant differences in COVID-19 hospitalizations were observed between self-reported white, Asian and black individuals (1-way ANOVA p<0.001). **B:** Mean COVID-19 hospitalizations per 10,000 individuals by deciles of the Townsend Deprivation Index. Townsend Deprivation Index was significantly associated with COVID-19 hospitalization in adjusted and unadjusted models (p<0.001). **C:** COVID-19 hospitalizations per 10,000 individuals by brackets of self-reported pre-tax household income. Pre-tax household income was significantly associated with COVID-19 hospitalization in an unadjusted model (p<0.001).

To understand the relationship between socioeconomic status and COVID-19 hospitalization, individuals were stratified at enrollment into deciles of the Townsend Deprivation Index, a previously validated composite metric based on employment status, car or home ownership, and household crowding.^5^ Participants with greater Townsend Deprivation Indices were at substantially higher risk of COVID-19 hospitalization, with a similar pattern observed based on self-reported household income (**Figure, Panels B, C**).

The relationship between race and COVID-19 hospitalization was only modestly attenuated in a logistic regression model that additionally adjusted for Townsend Deprivation Index and household income--odds ratios for black and Asian participants of 3.1 (95%CI 2.0–4.8) and 2.0 (95%CI 1.2–3.1) as compared to white participants respectively.

## DISCUSSION

Within a large population of participants in a national biobank, striking gradients in risk of hospitalization for COVID-19 were noted according to race and a metric of socioeconomic deprivation. The increased risk observed in black participants was attenuated but remained significant after adjusting for socioeconomic deprivation and household income. The extent to which this increased risk relates to variation in pre-existing cardiometabolic comorbidities, differences in testing or hospitalization patterns, or additional disparities in social determinants of health warrants further study.^6^ With respect to potential biologic factors, ongoing efforts seek to determine whether genetics--known to both vary substantially across racial groups and contribute to pre-existing comorbidities--plays an important role in COVID-19 disease severity.

This study has limitations. First, the UK Biobank enrolled individuals on a volunteer basis and is not a population-based study-additional efforts are needed to generalize these observations in other settings. Second, Townsend Deprivation Index and household income were assessed at enrollment, and participants’ status may have changed in subsequent years. Third, additional and more sophisticated analytic techniques may prove useful in further disentangling COVID-19 related disparities.

## Data Availability

Data is made available from the UK Biobank to researchers from universities and other research institutions with genuine research inquiries following IRB and UK Biobank approval.

## ACKNOWLEDGEMENTS

We are indebted to the UK Biobank and its participants who provided biological samples and data for this analysis. Work was performed under UK Biobank application #7089. Data analysis was approved by the Partners Healthcare institutional review board.

## REFERENCES

1. CDC. Coronavirus Disease 2019 (COVID-19). Centers for Disease Control and Prevention. https://www.cdc.gov/coronavirus/2019-ncov/need-extra-precautions/racial-ethnic-minorities.html. Published February 11, 2020. Accessed April 26, 2020.

2. Nearly a third of 200 blood samples taken in Chelsea show exposure to coronavirus - The Boston Globe. https://www.bostonglobe.com/2020/04/17/business/nearly-third-200-blood-samples-taken-chelsea-show-exposure-coronavirus/. Accessed April 26, 2020.

3. Bycroft C, Freeman C, Petkova D, et al. The UK Biobank resource with deep phenotyping and genomic data. Nature. 2018;562(7726):203–209. doi:10.1038/s41586-018-0579-z

4. Armstrong J, Rudkin J, Allen N, et al. Dynamic linkage of COVID-19 test results between Public Health England’s Second Generation Surveillance System and UK Biobank. 2020:2163001 Bytes. doi:10.6084/M9.FIGSHARE.12091455

5. Townsend P, Phillimore P, Beattie A. Health and Deprivation: Inequality and the North. London, England: Croom Helm; 1988.

6. Yancy CW. COVID-19 and African Americans. JAMA. April 2020. doi:10.1001/jama.2020.6548

